# Availability and prescribing of extended release buprenorphine injection for Medicaid beneficiaries, 2018-2022

**DOI:** 10.1101/2024.01.05.24300895

**Authors:** Rachael K. Ross, Kara E. Rudolph, Chelsea Shover

**Affiliations:** Department of Epidemiology, Mailman School of Public Health, Columbia University, New York, NY; Division of General Internal Medicine and Health Services Research, School of Medicine, University of California Los Angeles, Los Angeles, CA

**Keywords:** Extended release buprenorphine, injection buprenorphine, Sublocade, Brixadi, Medicaid

## Abstract

**Background and aims:** Extended release buprenorphine injection (INJ-BUP) has been available in the United States since 2018. INJ-BUP has the potential to positively impact OUD treatment outcomes by providing additional treatment options and patient choice. We aimed to describe the availability and prescribing of INJ-BUP for Medicaid beneficiaries since its availability, nationwide and by state.

**Methods:** To assess availability, we measured the number of substance use disorder (SUD) facilities that offered INJ-BUP and accepted Medicaid insurance in 2018-2021 and calculated the percentage of all facilities offering medications of OUD. To assess prescribing, we measured the number of prescriptions for INJ-BUP paid by Medicaid 2018-2022 and calculated the percentage of all buprenorphine prescriptions paid by Medicaid. Data sources were publicly available.

**Results:** The number of facilities that offered INJ-BUP and accepted Medicaid insurance increased from 360 (2.5% of all SUD facilities offering medication) in 2018 to 2,257 (13.3%) in 2021. The number of prescriptions for INJ-BUP paid by Medicaid increased from 4,322 (0.1% of all buprenorphine prescriptions) in 2018 to 186,861 (2.0%) in 2022. There was notable variability in the number of facilities and prescriptions by state.

**Conclusions:** There has been exponential increase in the number of INJ-BUP prescriptions but uptake is much less than observed in other countries in shorter time periods. Limited availability at SUD treatment facilities that accept Medicaid may be one barrier to access.

## INTRODUCTION

In 2017, a monthly buprenorphine injection (INJ-BUP), called Sublocade, was approved by the Food and Drug Administration for the treatment of opioid use disorder (OUD).(1) A second INJ-BUP, with both monthly and weekly dosages, called Brixadi, was approved in May 2023 and became available in September 2023.(2) These extended release formulations of buprenorphine have the potential to positively impact OUD treatment outcomes.(3,4) Compared to oral formulations, INJ-BUP has a greater initial medication dose and consistent bioavailability, reducing the risk of withdrawal symptoms.(5) There is no risk of misuse or diversion, reduced burden of daily medication, and, for some individuals, a periodic injection may reduce negative effects of stigma associated with opioid agonist treatment.(6–8)

Medicaid is the largest payer for behavioral health treatment, including OUD treatment, in the US; thus, Medicaid coverage is important for access and uptake of INJ-BUP. In 2018, INJ-BUP was not covered by Medicaid in 18 states according to a Substance Abuse and Mental Health Services Administration (SAMHSA) report; by 2022, at least 17 of those states reported some coverage (1 was unknown) in a Kaiser Family Foundation Survey of Medicaid Covered Behavioral Health Benefits for Adults.(9,10) Therefore Medicaid covered INJ-BUP in potentially all states within 4 years of approval. Availability of INJ-BUP at substance use disorder (SUD) treatment facilities and INJ-BUP prescriptions paid by Medicaid have not been described since 2019.(4,11) At that time there was limited availability of INJ-BUP at facilities that accepted Medicaid insurance. Here, we describe recent INJ-BUP availability and prescribing for Medicaid beneficiaries.

## METHODS

### Availability

We analyzed data from National Survey of Substance Abuse Treatment Services (N-SSATS) from 2017-2020(12) and the National Substance Use and Mental Health Services Survey (N-SUMHSS) from 2021.(13) These surveys, conducted by the SAMHSA, collect data on public and private substance abuse treatment facilities in the US. We restricted analysis to facilities that reported offering medications approved for OUD treatment including extended-release naltrexone, methadone, oral buprenorphine (with or with naloxone), INJ-BUP, and the buprenorphine implant (discontinued in 2020). We report the number and percentage of facilities that reported offering each route of buprenorphine and accepting Medicaid insurance, overall and by state. We also calculated the number of facilities per 1000 Medicaid beneficiaries treated for an OUD. For the number of beneficiaries treated for an OUD, we took the average of the numbers reported in the 2019 and 2020 Interactive T-MSIS SUD Date Book (from Table G.2 in the reference).(14)

### Prescribing

We analyzed State Drug Utilization Data (SDUD) from 2017-2022.(15) SDUD are reported by states for outpatient medications paid for by state Medicaid agencies. SDUD include the count of prescription fills for each National Drug Code (NDC) by calendar quarter, overall and by state. We identified buprenorphine prescriptions including route using a list of NDCs compiled from the Centers for Disease Control and Prevention’s Opioid NDC and Oral MME Conversion File 2017-2020 and the Centers for Medicare and Medicaid Services Chronic Conditions Data Warehouse definition for OUD. In SDUD, counts <11 are suppressed; we imputed suppressed counts with 5.5. We report the total count of prescription fills for each route of buprenorphine, overall and by state. We also calculated the number of prescriptions per 1000 Medicaid beneficiaries treated for an OUD (denominator described above).

## RESULTS

### Availability

The total number of substance abuse treatment facilities included in the N-SSATS and N-SUMHSS surveys that reported offering any medication for OUD treatment increased from 14,667 in 2018 (the first full year INJ-BUP was approved) to 17,333 in 2021 (Table 1). INJ-BUP was available at 360 facilities that accept Medicaid (2.5% of all SUD facilities offering medication) in 2018. That number increased steadily over the first four years 2,257 (13% of facilities) in 2021. In contrast, the percentage of facilities offering oral buprenorphine and accepting Medicaid increased from 23% to 36%. In 2021, for every 1000 Medicaid beneficiaries with OUD, there were approximately 3.9 facilities offering oral buprenorphine and 1.4 facilities offering INJ-BUP.

**Table 1.**
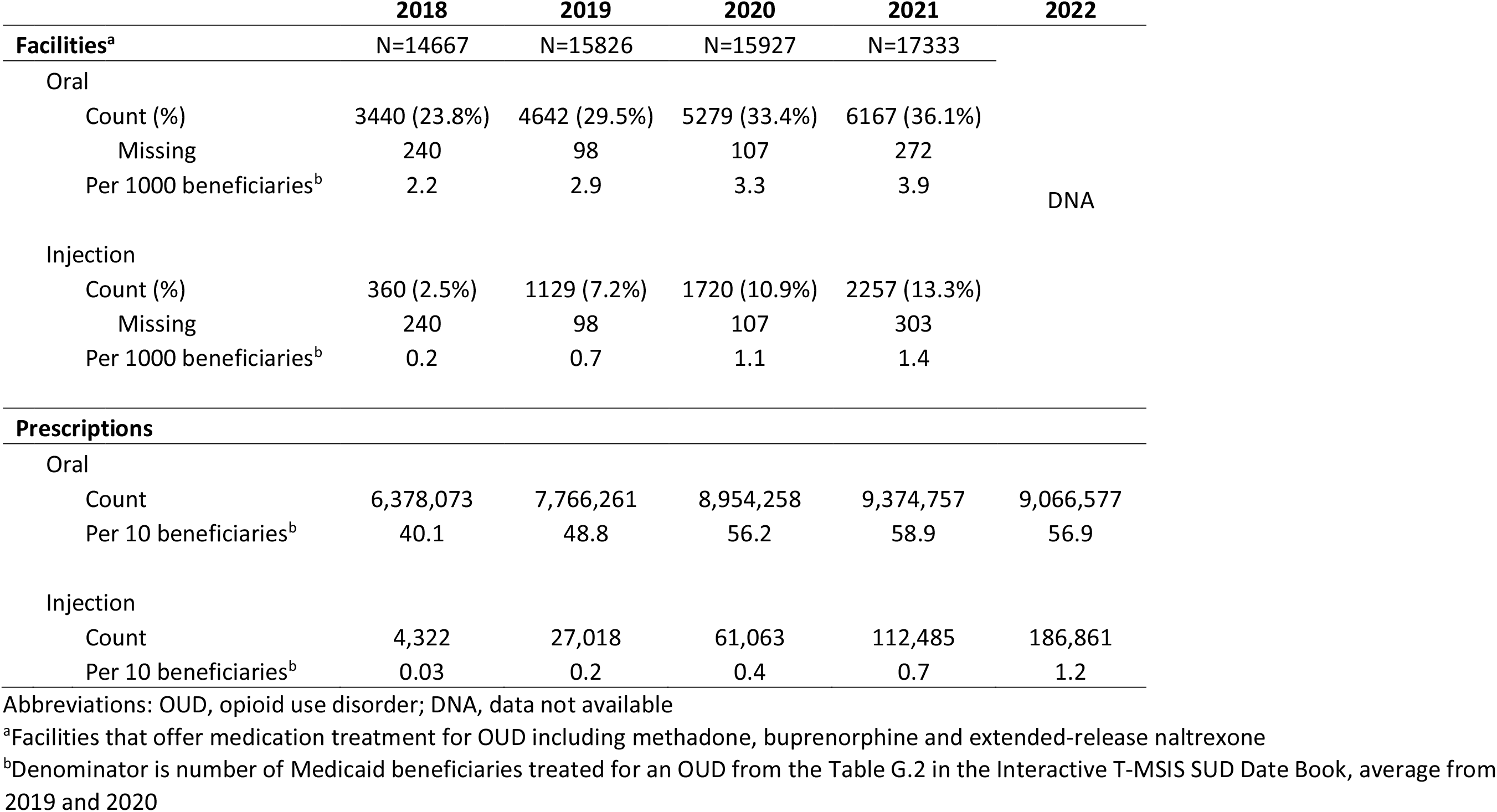
Availability of buprenorphine for Medicaid beneficiaries at substance use disorder treatment facilities offering medications for OUD treatment (top) and buprenorphine prescriptions for OUD treatment paid by Medicaid insurance (bottom)

|  | 2018 | 2019 | 2020 | 2021 | 2022 |
| --- | --- | --- | --- | --- | --- |
| Facilities<sup>a</sup> | N=14667 | N=15826 | N=15927 | N=17333 |  |
| Oral |  |  |  |  |  |
| Count (%) | 3440 (23.8%) | 4642 (29.5%) | 5279 (33.4%) | 6167 (36.1%) |  |
| Missing | 240 | 98 | 107 | 272 |  |
| Per 1000 beneficiaries<sup>b</sup> | 2.2 | 2.9 | 3.3 | 3.9 | DNA |
| Injection |  |  |  |  |  |
| Count (%) | 360 (2.5%) | 1129 (7.2%) | 1720 (10.9%) | 2257 (13.3%) |  |
| Missing | 240 | 98 | 107 | 303 |  |
| Per 1000 beneficiaries<sup>b</sup> | 0.2 | 0.7 | 1.1 | 1.4 |  |
| **Prescriptions** |  |  |  |  |  |
| Oral |  |  |  |  |  |
| Count | 6,378,073 | 7,766,261 | 8,954,258 | 9,374,757 | 9,066,577 |
| Per 10 beneficiaries<sup>b</sup> | 40.1 | 48.8 | 56.2 | 58.9 | 56.9 |
| Injection |  |  |  |  |  |
| Count | 4,322 | 27,018 | 61,063 | 112,485 | 186,861 |
| Per 10 beneficiaries<sup>b</sup> | 0.03 | 0.2 | 0.4 | 0.7 | 1.2 |

Abbreviations: OUD, opioid use disorder; DNA, data not available
<sup>a</sup>Facilities that offer medication treatment for OUD including methadone, buprenorphine and extended-release naltrexone
<sup>b</sup>Denominator is number of Medicaid beneficiaries treated for an OUD from the Table G.2 in the Interactive T-MSIS SUD Data Book, average from 2019 and 2020

Figure 1A depicts the number of facilities in each state by year (numbers provided in Appendix Table 1). In 2018, there were 5 states that did not have a single facility offering INJ-BUP and accepting Medicaid and most states (n=30) had <5 facilities. By 2019, there was just a single state (South Dakota) without any facilities and 7 states with <5. By 2020, all states had at least one facility. The median number of facilities offering INJ-BUP and accepting Medicaid in each state increased from 4 (interquartile range [IQR] 2-9) in 2018 to 29 (IQR 14-67) in 2021 (Table 2). Figure 1B depicts the percentage of facilities offering INJ-BUP and accepting Medicaid (of all SUD facilities offering medication) in each state by year (numbers provided in Appendix Table 2). The median percentage in each state increased from 1.9% (IQR 1.0-3.5) to 12.8 (IQR 8.5-18.9) (Table 2).

**Table 2.**
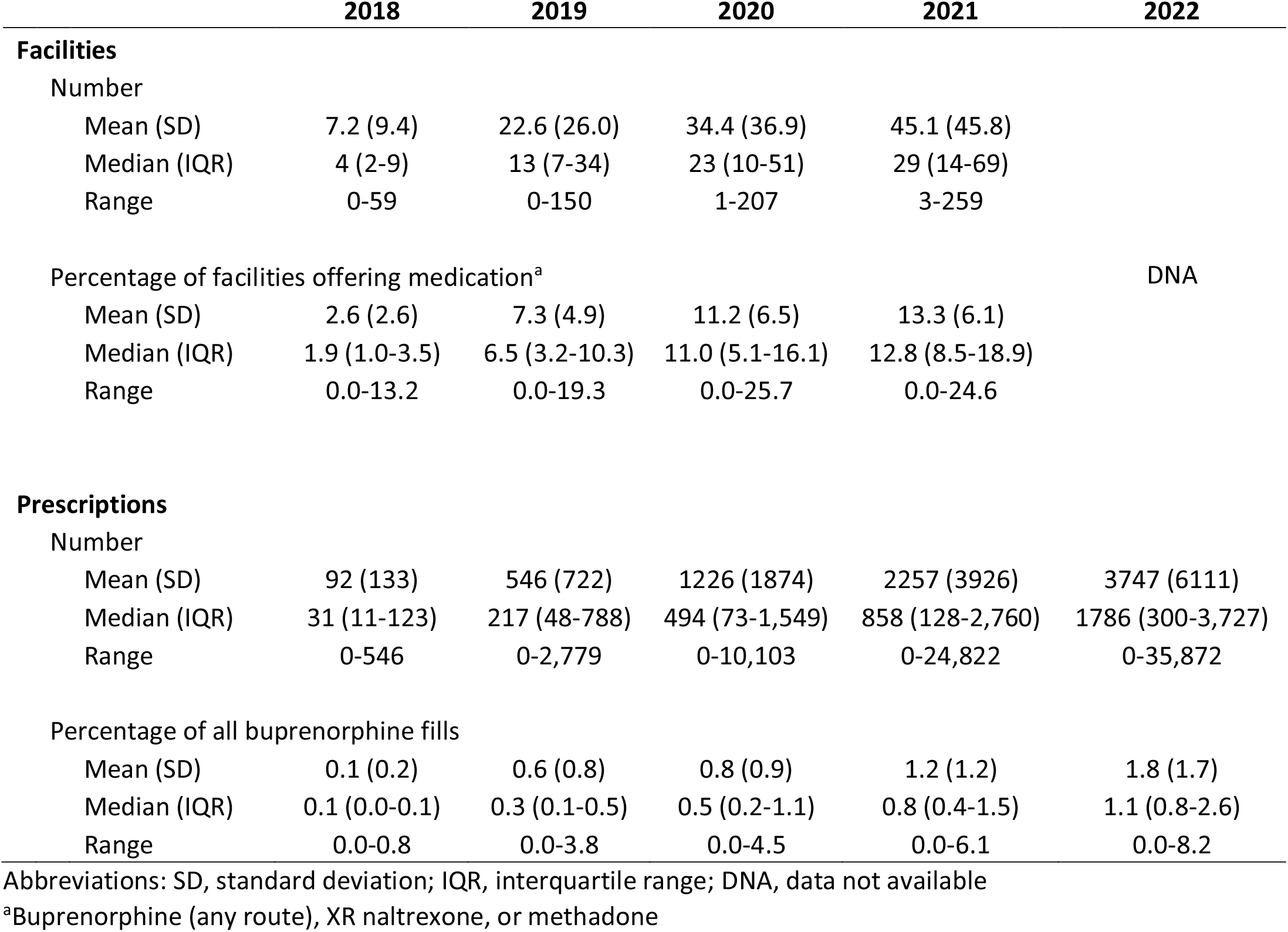
Summary statistics of the state-level number of facilities that accept Medicaid and offer buprenorphine injection and the number of prescriptions for buprenorphine injection paid by Medicaid.

**Figure 1.**
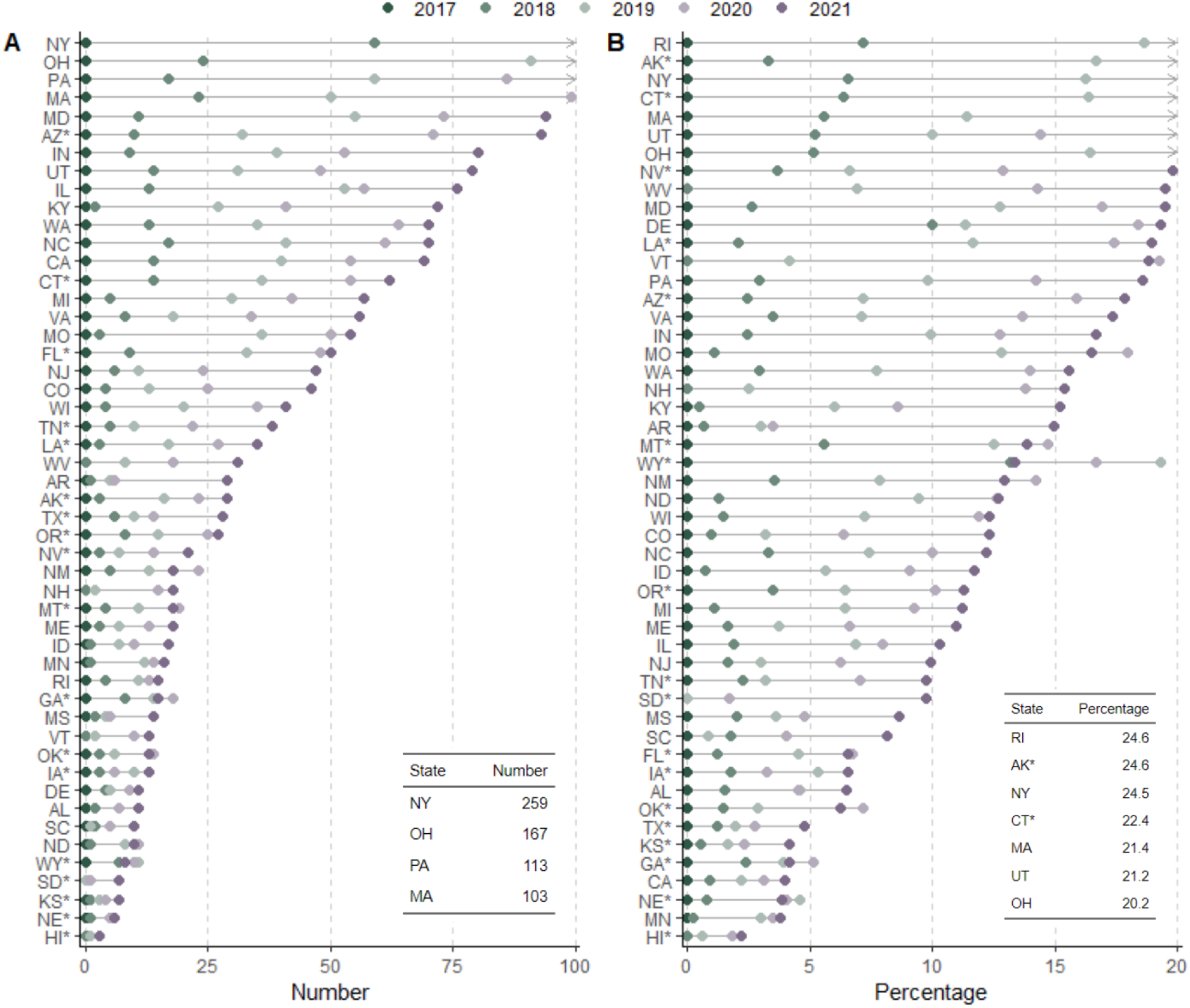
Substance use disorder treatment facilities that offered buprenorphine injection and accepted Medicaid insurance: absolute number (A) and as a percentage of all facilities offering medication for opioid use disorder (B). In each panel states are sorted by statistic in 2021. Arrows indicate that value exceeds the x-axis upper limit and are shown in the inset table. * marks states without Medicaid coverage for buprenorphine injection in 2018.

### Prescribing

Nationwide, there were 4,322 prescriptions for INJ-BUP paid by Medicaid in 2018, the first year it was available (Table 1). The number of prescriptions exponentially increased from 2018 to 2022. In 2022, there were 186,861 prescriptions. In contrast, the number of oral buprenorphine prescriptions increased from 6.4 million in 2018 to 9.1 million in 2022. In 2022, 2.0% of all buprenorphine prescriptions paid by Medicaid were INJ-BUP. In that year, there were approximately 1.2 prescriptions of INJ-BUP and 56.9 prescriptions of oral buprenorphine per 10 beneficiaries with OUD.

Figure 2A depicts the number of INJ-BUP prescriptions in each state by year (numbers provided in Appendix Table 1). In 2018, there were 13 states with more than 100 prescriptions for INJ-BUP. This number increased to 31 states in 2019, 36 states in 2020, and 40 states in 2021 and 2022. In 2022, 2 states did not report any prescriptions for INJ-BUP (Arkansas and Texas). The median number of prescriptions in each state went from 31 (IQR 11-107) in 2018 to 1786 (IQR 325-3609) in 2022 (Table 2). Figure 2B depicts the proportion of all buprenorphine prescriptions that were for INJ-BUP in each state by year (numbers provided in Appendix Table 2). The median percentage of prescriptions that were for INJ-BUP in each state increased from 0.1% (IQR 0.0%-0.1%) to 1.1% (IQR 0.8%-2.6%) (Table 2).

**Figure 2.**
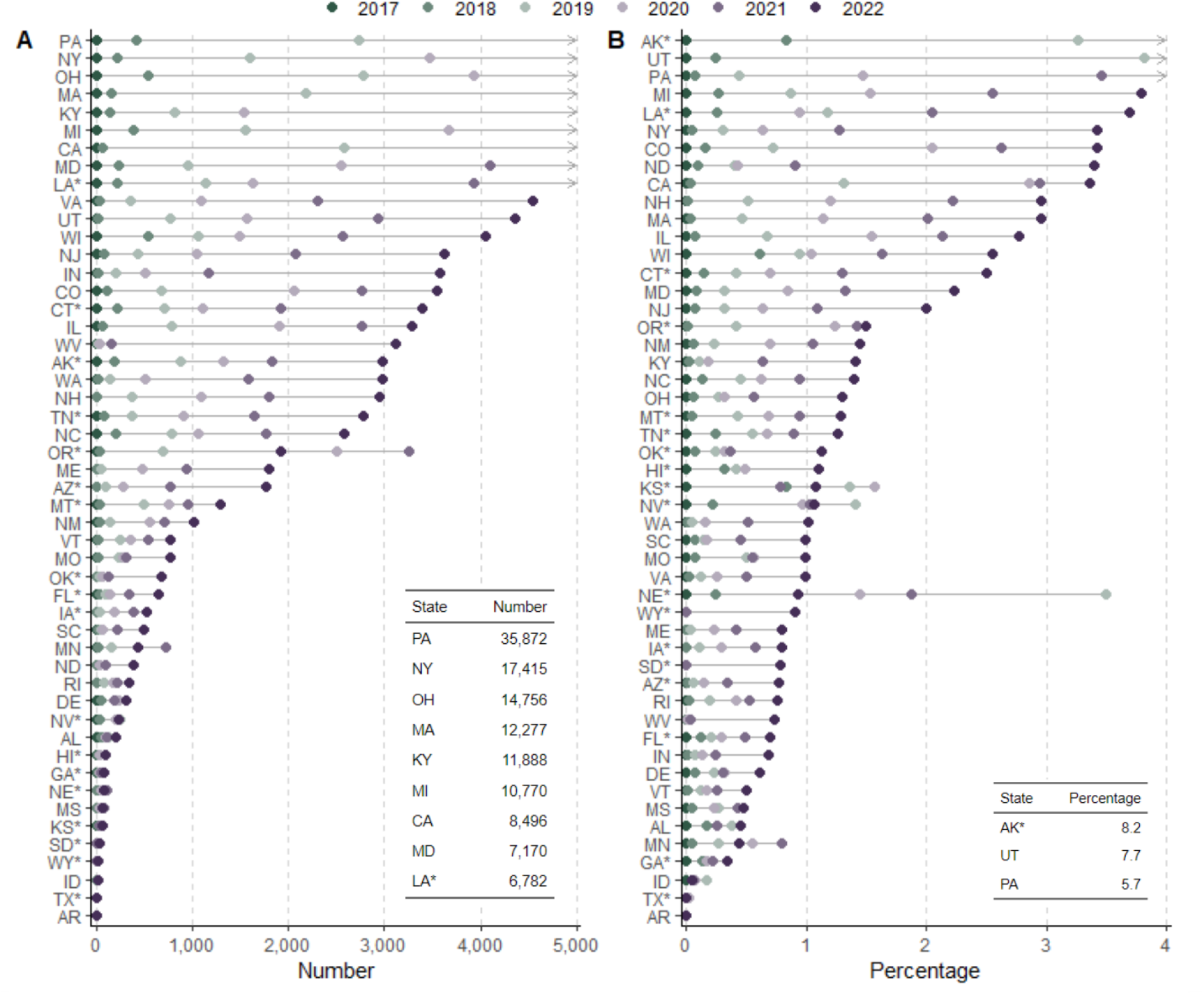
Prescription for buprenorphine injection paid by Medicaid: absolute number (A) and as a percentage of all buprenorphine prescriptions for opioid use disorder. In each panel states are sorted by statistic in 2022. Arrows indicate that value exceeds the x-axis upper limit and are shown in the inset table. * marks states without Medicaid coverage for buprenorphine injection in 2018.

## DISCUSSION

There has been notable uptake of INJ-BUP among Medicaid beneficiaries, particularly compared to the extended release buprenorphine implant previously.(4) However, INJ-BUP is still rare compared to oral buprenorphine. In contrast, in Australia, use of XR buprenorphine was as common as oral buprenorphine just over three years after it became available.(16) In Ontario, approximately 6% of people initiating opioid agonist treatment initiated XR buprenorphine just two years after approval.(17) In the US, limited availability at SUD treatment facilities that accept Medicaid may be a barrier to access. At present there is little information supporting treatment decisions between INJ-BUP and oral buprenorphine or another MOUD such as XR naltrexone,(18) though research suggests that increased treatment options and patient choice can improve treatment adherence and outcomes.(19) Limitations of this work include lack of data on access to INJ-BUP in other healthcare settings and INJ-BUP provided in the office settings without a pharmacy fill; both limitations suggest that our results underestimate availability and prescribing. Characterizing and ultimately eliminating barriers to INJ-BUP access are key to leveraging this novel modality to improve OUD outcomes.

## Data Availability

The data that support the findings of this study are openly available at https://www.samhsa.gov/data/data-we-collect/n-ssats-national-survey-substance-abuse-treatment-services, https://data.medicaid.gov/datasets?theme%5B0%5D=State+Drug+Utilization, and https://portal.cms.gov/MSTR2021/servlet/mstrWeb?src=mstrWeb.2048001&ru=1&documentID=E8E899A911EB08AB795A0080EFE5D251. Code for this analysis is available at https://github.com/rachael-k-ross/Inj-BUP-Availability-Prescribing.

## References

1. Poliwoda S, Noor N, Jenkins JS, Stark CW, Steib M, Hasoon J, et al. Buprenorphine and its formulations: a comprehensive review. Health Psychol Res [Internet]. 2022 Aug 20 [cited 2023 Aug 16];10(3). Available from: https://healthpsychologyresearch.openmedicalpublishing.org/article/37517-buprenorphine-and-its-formulations-a-comprehensive-review

2. braeburn [Internet]. [cited 2023 Oct 7]. BRIXADI® (buprenorphine) Extended-Release Injection for Subcutaneous Use (CIII) is Now Available in the U.S. for the Treatment of Moderate to Severe Opioid Use Disorder. Available from: https://braeburnrx.com/brixadi-buprenorphine-extended-release-injection-for-subcutaneous-use-ciii-is-now-available-in-the-u-s-for-the-treatment-of-moderate-to-severe-opioid-use-disorder/

3. Maremmani I, Dematteis M, Gorzelanczyk EJ, Mugelli A, Walcher S, Torrens M. Long-Acting Buprenorphine Formulations as a New Strategy for the Treatment of Opioid Use Disorder. J Clin Med. 2023 Aug 26;12(17):5575.

4. Shover CL. Commentary on Larance et al. (2020): Priorities and concerns of people who use opioids are key to scaling up XR-buprenorphine. Addiction. 2020 Jul;115(7):1306–7.

5. Roberts E, Humphreys K. Does the advent of depot therapy represent a step change in our understanding of opioid use disorder and its treatment? Drug Alcohol Rev. 2023 Jul 31;dar.13732.

6. Treloar C, Lancaster K, Gendera S, Rhodes T, Shahbazi J, Byrne M, et al. Can a new formulation of opiate agonist treatment alter stigma?: Place, time and things in the experience of extended-release buprenorphine depot. Int J Drug Policy. 2022 Sep;107:103788.

7. Somaini L, Vecchio S, Corte C, Coppola C, Mahony A, Pitts A, et al. Prolonged-Release Buprenorphine Therapy in Opioid Use Disorder Can Address Stigma and Improve Patient Quality of Life. Cureus [Internet]. 2021 Oct 5 [cited 2023 Aug 22]; Available from: https://www.cureus.com/articles/72443-prolonged-release-buprenorphine-therapy-in-opioid-use-disorder-can-address-stigma-and-improve-patient-quality-of-life

8. Gilman M, Li L, Hudson K, Lumley T, Myers G, Corte C, et al. Current and future options for opioid use disorder: a survey assessing real-world opinion of service users on novel therapies including depot formulations of buprenorphine. Patient Prefer Adherence. 2018 Oct;Volume 12:2123–9.

9. O’Brien P, Alikhan S, Cummings N, Hohlbauch A, Hughey L, Schrader K, et al. Medicaid Coverage of Medication-Assisted Treatment for Alcohol and Opioid Use Disorders and of Medication for the Reversal of Opioid Overdose [Internet]. Subtance Abuse and Mental Health Services Administration; 2018 [cited 2023 Oct 7]. Report No.: SMA-18-5093. Available from: https://store.samhsa.gov/sites/default/files/d7/priv/medicaidfinancingmatreport_0.pdf

10. Kaiser Family Foundation [Internet]. [cited 2023 Oct 4]. Medicaid Behavioral Health Services: Injectable Buprenorphine for Medication Assisted Treatment (MAT). Available from: https://www.kff.org/other/state-indicator/medicaid-behavioral-health-services-injectable-buprenorphine-for-medication-assisted-treatment-mat/

11. Shover CL. Availability of Extended-Release Buprenorphine to Treat Opioid Use Disorders Among Medicaid-Covered Patients. Psychiatr Serv. 2021 Feb 1;72(2):225–6.

12. Substance Abuse and Mental Health Services Administration [Internet]. [cited 2023 Sep 13]. National Survey of Substance Abuse Treatment Services (N-SSATS). Available from: https://www.samhsa.gov/data/data-we-collect/n-ssats-national-survey-substance-abuse-treatment-services

13. Substance Abuse and Mental Health Services Administration [Internet]. [cited 2023 Sep 13]. National Substance Use and Mental Health Services Survey (N-SUMHSS). Available from: https://www.samhsa.gov/data/data-we-collect/n-sumhss-national-substance-use-and-mental-

14. health-services-survey Interactive T-MSIS SUD Data Book [Internet]. [cited 2023 Sep 15]. Available from: https://portal.cms.gov/MSTR2021/servlet/mstrWeb?src=mstrWeb.2048001=ru=1=documentID=E8E899A911EB08AB795A0080EFE5D251=evt=2048001=share=1=hiddensections=header%2Cpath%2CdockTop%2CdockLeft%2Cfooter=Server=V343069P=Port=0=Project=SUD+Data+Book_Prd=

15. Data.Medicaid.gov [Internet]. [cited 2023 Oct 4]. State Drug Utilization Data. Available from: https://data.medicaid.gov/datasets?theme%5B0%5D=State+Drug+Utilization

16. Bharat C, Chidwick K, Gisev N, Farrell M, Ali R, Degenhardt L. Trends in use of medicines for opioid agonist treatment in Australia, 2013–2022. Int J Drug Policy. 2024 Jan;123:104255.

17. Iacono A, Wang T, Tadrous M, Campbell T, Kolla G, Leece P, et al. Characteristics, treatment patterns and retention with extended-release subcutaneous buprenorphine for opioid use disorder: a population-based cohort study in Ontario, Canada. Drug Alcohol Depend. 2023 Nov;111032.

18. The ASAM National Practice Guideline for the Treatment of Opioid Use Disorder: 2020 Focused Update. J Addict Med. 2020 Mar;14(2S):1–91.

19. Yarborough BJH, Stumbo SP, McCarty D, Mertens J, Weisner C, Green CA. Methadone, buprenorphine and preferences for opioid agonist treatment: A qualitative analysis. Drug Alcohol Depend. 2016 Mar;160:112–8.

