## Appendix for "Availability and prescribing of extended release buprenorphine injection for Medicaid beneficiaries, 2018-2022"

Contents

Appendix Table 1. Number of facilities and prescriptions by state

Appendix Table 2. Percentage of facilities and prescriptions by state

**Appendix Table 1**. Number of substance use disorder facilities that offered injection buprenorphine and accepted Medicaid (left) and number of prescriptions for injection buprenorphine paid by Medicaid (right)

| | Facilities | | | | Prescriptions | | | | |
|---|---|---|---|---|---|---|---|---|---|
| **State** | 2018 | 2019 | 2020 | 2021 | 2018 | 2019 | 2020 | 2021 | 2022 |
| US | 360 | 1,129 | 1,720 | 2,257 | 4,322 | 27,018 | 61,063 | 112,485 | 186,861 |
| AK | 3 | 16 | 23 | 29 | 196 | 882 | 1,329 | 1,832 | 2,986 |
| AL | 2 | 7 | 7 | 11 | 53 | 134 | 104 | 117 | 206 |
| AR | 1 | 5 | 6 | 29 | 0 | 0 | 0 | 0 | 0 |
| AZ | 10 | 32 | 71 | 93 | 11 | 99 | 290 | 778 | 1,775 |
| CA | 14 | 40 | 54 | 69 | 63 | 2,576 | 6,240 | 7,114 | 8,497 |
| CO | 4 | 13 | 25 | 46 | 114 | 678 | 2,058 | 2,760 | 3,547 |
| CT | 14 | 36 | 54 | 62 | 220 | 710 | 1,105 | 1,916 | 3,399 |
| DE | 4 | 5 | 9 | 11 | 51 | 193 | 231 | 196 | 319 |
| FL | 9 | 33 | 48 | 50 | 44 | 96 | 152 | 350 | 654 |
| GA | 8 | 14 | 18 | 15 | 22 | 33 | 39 | 53 | 83 |
| HI | 0 | 1 | 3 | 3 | 28 | 33 | 44 | 98 | 98 |
| IA | 3 | 10 | 6 | 13 | 0 | 33 | 187 | 384 | 536 |
| ID | 1 | 7 | 10 | 17 | 6 | 17 | 17 | 17 | 17 |
| IL | 13 | 53 | 57 | 76 | 70 | 790 | 1,903 | 2,762 | 3,287 |
| IN | 9 | 39 | 53 | 80 | 28 | 202 | 508 | 1,168 | 3,579 |
| KS | 1 | 3 | 4 | 7 | 28 | 50 | 72 | 44 | 67 |
| KY | 2 | 27 | 41 | 72 | 148 | 822 | 1,542 | 5,521 | 11,888 |
| LA | 3 | 17 | 27 | 35 | 216 | 1,145 | 1,638 | 3,936 | 6,782 |
| MA | 23 | 50 | 99 | 103 | 162 | 2,189 | 5,254 | 9,003 | 12,277 |
| MD | 11 | 55 | 73 | 94 | 232 | 951 | 2,545 | 4,103 | 7,170 |
| ME | 3 | 7 | 13 | 18 | 0 | 51 | 480 | 937 | 1,797 |
| MI | 5 | 30 | 42 | 57 | 387 | 1,557 | 3,668 | 6,900 | 10,771 |
| MN | 1 | 12 | 14 | 16 | 22 | 162 | 437 | 729 | 438 |
| MO | 3 | 36 | 50 | 54 | 29 | 232 | 284 | 316 | 766 |
| MS | 2 | 4 | 5 | 14 | 6 | 39 | 39 | 79 | 67 |
| MT | 4 | 11 | 19 | 18 | 39 | 504 | 755 | 962 | 1,301 |
| NC | 17 | 41 | 61 | 70 | 212 | 787 | 1,063 | 1,768 | 2,581 |
| ND | 1 | 8 | 11 | 10 | 6 | 33 | 39 | 95 | 385 |
| NE | 1 | 6 | 5 | 6 | 6 | 93 | 50 | 120 | 79 |
| NH | 0 | 2 | 15 | 18 | 6 | 372 | 1,095 | 1,798 | 2,949 |
| NJ | 6 | 11 | 24 | 47 | 87 | 432 | 1,045 | 2,081 | 3,619 |
| NM | 5 | 13 | 23 | 18 | 42 | 151 | 566 | 710 | 1,017 |
| NV | 3 | 7 | 14 | 21 | 33 | 249 | 204 | 240 | 244 |
| NY | 59 | 150 | 207 | 259 | 226 | 1,600 | 3,478 | 6,808 | 17,415 |
| OH | 24 | 91 | 130 | 167 | 537 | 2,779 | 3,935 | 6,759 | 14,756 |
| OK | 3 | 6 | 14 | 13 | 11 | 44 | 74 | 131 | 686 |
| OR | 8 | 15 | 25 | 27 | 35 | 690 | 2,499 | 3,263 | 1,925 |
| PA | 17 | 59 | 86 | 113 | 424 | 2,733 | 10,103 | 24,822 | 35,872 |
| RI | 4 | 11 | 13 | 15 | 11 | 91 | 180 | 229 | 343 |
| SC | 2 | 1 | 5 | 10 | 22 | 59 | 76 | 219 | 494 |
| SD | 0 | 0 | 1 | 7 | 0 | 0 | 0 | 0 | 39 |
| TN | 5 | 10 | 22 | 38 | 81 | 368 | 917 | 1,651 | 2,784 |
| TX | 6 | 10 | 14 | 28 | 0 | 0 | 6 | 0 | 0 |
| UT | 14 | 31 | 48 | 79 | 28 | 778 | 1,571 | 2,939 | 4,367 |
| VA | 8 | 18 | 34 | 56 | 33 | 360 | 1,090 | 2,304 | 4,542 |
| VT | 0 | 2 | 10 | 13 | 25 | 250 | 354 | 550 | 775 |
| WA | 13 | 35 | 64 | 70 | 26 | 147 | 507 | 1,579 | 2,983 |
| WI | 4 | 20 | 35 | 41 | 546 | 1,071 | 1,498 | 2,569 | 4,051 |
| WV | 0 | 8 | 18 | 31 | 17 | 28 | 39 | 156 | 3,112 |
| WY | 7 | 11 | 10 | 8 | 0 | 0 | 0 | 0 | 22 |

**Appendix Table 2**. Percentage of substance use disorder facilities that offered injection buprenorphine and accepted Medicaid (left) and percentage of buprenorphine prescriptions paid by Medicaid that were injection (right)

| | Facilities[a] | | | | Prescriptions | | | | |
|---|---|---|---|---|---|---|---|---|---|
| **State** | 2018 | 2019 | 2020 | 2021 | 2018 | 2019 | 2020 | 2021 | 2022 |
| US | 2% | 7% | 11% | 13% | 0.1% | 0.3% | 0.7% | 1.2% | 2.0% |
| AK | 3% | 17% | 22% | 25% | 0.8% | 3.3% | 4.5% | 5.4% | 8.2% |
| AL | 2% | 5% | 5% | 6% | 0.2% | 0.4% | 0.3% | 0.3% | 0.4% |
| AR | 1% | 3% | 4% | 15% | 0.0% | 0.0% | 0.0% | 0.0% | 0.0% |
| AZ | 2% | 7% | 16% | 18% | 0.0% | 0.1% | 0.1% | 0.3% | 0.8% |
| CA | 1% | 2% | 3% | 4% | 0.0% | 1.3% | 2.9% | 2.9% | 3.4% |
| CO | 1% | 3% | 6% | 12% | 0.2% | 0.7% | 2.0% | 2.6% | 3.4% |
| CT | 6% | 16% | 26% | 22% | 0.1% | 0.4% | 0.7% | 1.3% | 2.5% |
| DE | 10% | 11% | 18% | 19% | 0.1% | 0.2% | 0.3% | 0.3% | 0.6% |
| FL | 1% | 5% | 7% | 7% | 0.1% | 0.2% | 0.3% | 0.5% | 0.7% |
| GA | 2% | 4% | 5% | 4% | 0.1% | 0.2% | 0.2% | 0.2% | 0.3% |
| HI | 0% | 1% | 2% | 2% | 0.3% | 0.4% | 0.5% | 1.1% | 1.1% |
| IA | 2% | 5% | 3% | 7% | 0.0% | 0.1% | 0.3% | 0.6% | 0.8% |
| ID | 1% | 6% | 9% | 12% | 0.1% | 0.2% | 0.1% | 0.1% | 0.0% |
| IL | 2% | 7% | 8% | 10% | 0.1% | 0.7% | 1.5% | 2.1% | 2.8% |
| IN | 2% | 10% | 13% | 17% | 0.0% | 0.1% | 0.1% | 0.2% | 0.7% |
| KS | 1% | 2% | 2% | 4% | 0.8% | 1.4% | 1.6% | 0.8% | 1.1% |
| KY | 0% | 6% | 9% | 15% | 0.0% | 0.1% | 0.2% | 0.6% | 1.4% |
| LA | 2% | 12% | 17% | 19% | 0.3% | 1.2% | 0.9% | 2.1% | 3.7% |
| MA | 6% | 11% | 23% | 21% | 0.0% | 0.5% | 1.1% | 2.0% | 3.0% |
| MD | 3% | 13% | 17% | 19% | 0.1% | 0.3% | 0.8% | 1.3% | 2.2% |
| ME | 2% | 4% | 7% | 11% | 0.0% | 0.0% | 0.2% | 0.4% | 0.8% |
| MI | 1% | 6% | 9% | 11% | 0.3% | 0.9% | 1.5% | 2.6% | 3.8% |
| MN | 0% | 3% | 3% | 4% | 0.0% | 0.3% | 0.6% | 0.8% | 0.4% |
| MO | 1% | 13% | 18% | 17% | 0.1% | 0.5% | 0.6% | 0.6% | 1.0% |
| MS | 2% | 4% | 5% | 9% | 0.0% | 0.3% | 0.2% | 0.4% | 0.5% |
| MT | 6% | 13% | 15% | 14% | 0.0% | 0.4% | 0.7% | 0.9% | 1.3% |
| NC | 3% | 7% | 10% | 12% | 0.1% | 0.5% | 0.6% | 0.9% | 1.4% |
| ND | 1% | 9% | 13% | 13% | 0.1% | 0.4% | 0.4% | 0.9% | 3.4% |
| NE | 1% | 5% | 4% | 4% | 0.2% | 3.5% | 1.5% | 1.9% | 0.9% |
| NH | 0% | 3% | 14% | 15% | 0.0% | 0.5% | 1.2% | 2.2% | 3.0% |
| NJ | 2% | 3% | 6% | 10% | 0.1% | 0.3% | 0.6% | 1.1% | 2.0% |
| NM | 4% | 8% | 14% | 13% | 0.1% | 0.2% | 0.7% | 1.1% | 1.4% |
| NV | 4% | 7% | 13% | 20% | 0.2% | 1.4% | 1.0% | 1.0% | 1.1% |
| NY | 7% | 16% | 23% | 25% | 0.0% | 0.3% | 0.6% | 1.3% | 3.4% |
| OH | 5% | 16% | 21% | 20% | 0.1% | 0.3% | 0.3% | 0.6% | 1.3% |
| OK | 2% | 3% | 7% | 6% | 0.1% | 0.2% | 0.3% | 0.4% | 1.1% |
| OR | 3% | 6% | 10% | 11% | 0.0% | 0.4% | 1.2% | 1.4% | 1.5% |
| PA | 3% | 10% | 14% | 19% | 0.1% | 0.4% | 1.5% | 3.5% | 5.7% |
| RI | 7% | 19% | 21% | 25% | 0.0% | 0.2% | 0.4% | 0.5% | 0.8% |
| SC | 2% | 1% | 4% | 8% | 0.1% | 0.1% | 0.2% | 0.5% | 1.0% |
| SD | 0% | 0% | 2% | 10% | 0.0% | 0.0% | 0.0% | 0.0% | 0.8% |
| TN | 2% | 3% | 7% | 10% | 0.2% | 0.6% | 0.7% | 0.9% | 1.3% |
| TX | 1% | 2% | 3% | 5% | 0.0% | 0.0% | 0.0% | 0.0% | 0.0% |
| UT | 5% | 10% | 14% | 21% | 0.2% | 3.8% | 4.4% | 6.1% | 7.7% |
| VA | 3% | 7% | 14% | 17% | 0.0% | 0.1% | 0.3% | 0.5% | 1.0% |
| VT | 0% | 4% | 19% | 19% | 0.0% | 0.1% | 0.2% | 0.3% | 0.5% |
| WA | 3% | 8% | 14% | 16% | 0.0% | 0.0% | 0.2% | 0.5% | 1.0% |
| WI | 1% | 7% | 12% | 12% | 0.6% | 0.9% | 1.0% | 1.6% | 2.6% |
| WV | 0% | 7% | 14% | 19% | 0.0% | 0.0% | 0.0% | 0.0% | 0.7% |
| WY | 13% | 19% | 17% | 13% | 0.0% | 0.0% | 0.0% | 0.0% | 0.9% |

[a]Among facilities that offered medication for opioid use disorder
